# Comparative effectiveness of the monovalent XBB.1.5-containing covid-19 mRNA vaccine across three Nordic countries

**DOI:** 10.1101/2024.05.08.24307058

**Authors:** Niklas Worm Andersson, Emilia Myrup Thiesson, Nicklas Pihlström, Jori Perälä, Kristýna Faksová, Mie Agermose Gram, Eero Poukka, Tuija Leino, Rickard Ljung, Anders Hviid

**Affiliations:** Department of Epidemiology Research, Statens Serum Institut, Copenhagen, Denmark; Division of Use and Information, Swedish Medical Products Agency, Uppsala, Sweden; Institute of Environmental Medicine, Karolinska Institutet, Stockholm, Sweden; Infectious Disease Control and Vaccinations Unit, Department of Health Security, Finnish Institute for Health and Welfare, Helsinki, Finland; Department of Public Health, Faculty of Medicine, University of Helsinki, Helsinki, Finland; Pharmacovigilance Research Center, Department of Drug Design and Pharmacology, Faculty of Health and Medical Sciences, University of Copenhagen, Copenhagen, Denmark

## Abstract

**Objective:** To estimate the effectiveness of vaccination with a monovalent XBB.1.5-containing covid-19 mRNA vaccine against severe covid-19 across three Nordic countries.

**Design:** Nationwide cohort studies, using target trial emulation.

**Setting:** Denmark, Finland, and Sweden, from 1 October 2023 to 29 February 2024.

**Participants:** Individuals aged ≥65 years who had previously received at least four covid-19 vaccine doses. **Main outcome measures:** Cumulative incidences of covid-19 hospital admission and death for 12 weeks after immunisation (defined as 1 week after vaccination) among recipients of an XBB.1.5-containing covid-19 mRNA vaccine and matched non-recipients. Cumulative incidences were used to calculate comparative vaccine effectiveness (1-risk ratio) and risk differences.

**Results:** During autumn and winter 2023-2024, a total of 1,867,448 1:1 matched pairs of XBB-containing covid-19 mRNA vaccine recipients and non-recipients were included (mean age 75.4 years, standard deviation 7.4 years). The comparative vaccine effectiveness was 60.6% (95% confidence interval, 46.1% to 75.1%) against covid-19 hospital admission (930 *v* 2,551 events) and 77.9% (69.2% to 86.7%) against covid-19 related death (301 *v* 1,326 events) at 12 weeks of follow-up. This corresponded to 191.1 (95% confidence interval, 50.2 to 332.1) covid-19 hospital admissions and 109.2 (100.2 to 118.1) deaths prevented per 100,000 individuals vaccinated with an XBB.1.5-containing vaccine. The comparative vaccine effectiveness was similar across sex, age (65-74/≥75 years), number of previous covid-19 vaccine doses received, and seasonal influenza vaccination co-administration subgroups and periods of either omicron XBB- or BA.2.86-sublineage dominance. While the protection was highest during the first weeks after vaccination, it was well-preserved at end of week 12 of follow-up.

**Conclusion:** Among adults aged ≥65 years, vaccination with a monovalent XBB.1.5-containing covid-19 mRNA vaccine reduced the rates of covid-19 related hospital admission and death during autumn and winter 2023-2024 across three Nordic countries.

## INTRODUCTION

The monovalent omicron XBB.1.5-containing covid-19 mRNA vaccine was authorised in Europe and the US and implemented in the autumn and winter 2023-2024 covid-19 vaccination programs.[1,2] In Denmark, Finland, and Sweden, the XBB.1.5-containing mRNA vaccine was recommended as an additional covid-19 vaccine dose to individuals of the general population aged ≥65 years from 1 October 2023.

Clinical studies have shown that the XBB.1.5-containing mRNA vaccine is immunogenic against the autumn and winter 2023-2024-season prevailing omicron subvariants including both the XBB- and BA.2.86-sublineages (e.g., EG.5.1 and JN.1, respectively).[3,4] Evaluations of the vaccine effectiveness with respect to the prevention of severe covid-19, however, are rare and mainly reflects early-season short-term effectiveness with little follow-up.[5–8]

Across the three Nordic countries of Denmark, Finland, and Sweden, we estimated the comparative effectiveness of the monovalent XBB.1.5-containing covid-19 mRNA vaccine against hospital admission and death related to covid-19 in nationwide cohort analysis of adults aged ≥65 years with 12 weeks of follow-up.

## METHODS

### Study data sources, design, and cohort

In all three countries, we linked demography and healthcare data across different nationwide registries by using the country-specific unique identifiers assigned to all residents. Hereby, we retrieved individual-level information on covid-19 vaccinations, hospital admissions and recorded disease diagnoses, laboratory confirmed severe acute respiratory syndrome coronavirus 2 (SARS-CoV-2) infections, and demographic (age, sex, residency, healthcare occupation, and vital status) variables (see supplementary tables S1-S2 for further details).

We designed this non-interventional study building on the target trial emulation framework. Specifically, we compared the rates of hospital admission and death related to covid-19 according to receiving or not receiving the XBB.1.5-containing vaccine as an additional covid-19 dose during the study period 1 October 2023 to 29 February 2024 (supplementary table S3 lists the key components of such pragmatic target trial specification and emulation).[9,10] Across the three Nordic countries, the XBB-sublineage (particularly EG.5.1) was predominating until the end of November 2023 from where the BA.2.86-sublineage (particularly JN.1) predominated the remaining study period; the autumn and winter 2023-2024 covid-19 wave peaked around mid-November till mid-December 2023.

See supplementary text for a description of ethical approvals/exemptions.

We specified the following eligibility criteria which were assessed at start of the study period: aged ≥65 years, having country residency (to ensure a linkable identifier), no prior hospital admission related to covid-19, and prior receipt of ≥four covid-19 vaccine doses (of AZD1222, BNT162b2, or mRNA-1273 vaccines only [AZD1222 as part of the primary vaccination course only]); to construct a cohort representative of the general population targeted for vaccination with the XBB.1.5-containing vaccine during autumn and winter 2023-2024 as per the national covid-19 vaccination strategies.

### Outcomes

We defined covid-19 related hospital admission as any first inpatient hospital admission with a registered covid-19 related diagnosis and a positive polymerase chain reaction (PCR) test for SARS-CoV-2 (tested positive within 14 days before to two days after the day of admission) and covid-19 related death as any death within 30 days of a positive PCR test for SARS-CoV-2. The two outcomes were studied separately; the day of admission or death served as the respective event date. See supplementary table S4 for additional outcome definition details.

### Procedures

Individuals receiving an XBB.1.5-containing vaccine dose during the study period were matched on the day of vaccination with individuals who had not received an additional dose up until and including this day. We matched XBB.1.5-containing vaccine recipients and non-recipients 1:1 by exact matching without replacement on age (in 5-year bins), calendar time of last prior covid-19 vaccine dose received (in monthly bins; e.g., month of receiving the 4th dose for matched pairs where the XBB.1.5-containing vaccine was administered as a 5th dose), sex, region of residence, vaccination priority groups (i.e., individuals deemed at high-risk of severe covid-19 and healthcare personnel), and number of selected comorbidities (by 0, 1, 2, or ≥3 of chronic pulmonary disease, cardiovascular conditions, diabetes, autoimmunity-related conditions, cancer, and moderate-to-severe renal disease) (supplementary table S2) by number of prior covid-19 vaccine doses received.

The day the XBB.1.5-containing vaccine was administered within each matched pair served as the index date for both individuals. We followed individuals from 1 week after the index date (i.e., day 8; to ensure full immunisation among XBB.1.5-containing vaccine recipients) for outcome events until 12 weeks of follow-up had passed (that is, 91 days since the index date), receipt of an additional covid-19 vaccine dose, death, emigration, or end of the study period (29 February 2024), whichever occurred first. Additionally, if individuals who were included as a matched non-recipient received a covid-19 vaccine later than the assigned index date, follow-up was censored for the current matched pair and the now vaccinated individual could potentially re-enter the study as an XBB.1.5-containing vaccine recipient in a new matched pair on that given date if successfully matched to another non-recipient. Supplementary figure S1 shows a graphical illustration of our study design. At the time of running the analyses, data on covid-19 related hospitalisation in Sweden was only available until 31 December 2023; therefore, we ended follow-up on day 87 in Sweden for this specific analysis.

### Statistical analysis

We used the Aalen-Johansen estimator to obtain cumulative incidences of the outcomes among XBB-1.5 containing vaccine recipients and non-recipients; any death and non-covid-19 related death served as a competing risk for the covid-19 related hospital admission and covid-19 related death outcome analysis, respectively. Relative (that is, comparative vaccine effectiveness; calculated as 1–risk ratio) and absolute (that is, the estimated number of outcome events prevented by vaccination; reported per 100,000 individuals) risk differences were calculated from the cumulative incidences at 12-week follow-up. The corresponding 95% confidence intervals were calculated using the delta method; upper 95% confidence intervals for the comparative vaccine effectiveness estimates were truncated at 100% if higher. We combined country-specific estimates by random-effects meta-analyses implemented using the *mixmeta* package in R, which allows for potential variation in effect across countries.[11] Counts smaller than five could not be reported owing to privacy regulations while zero could be reported.

Subgroup analyses were done by sex (female/male), age groups (64-75/≥75 years), number of previous covid-19 vaccine doses (i.e., the XBB.1.5-containing vaccine received as the fifth, sixth, or seventh dose; ≥eighth dose was too few for separate analysis), and seasonal influenza vaccination: 1) co-administered on the same date, 2) received influenza vaccine within 1 week before to 1 week after the XBB.1.5-containing vaccine but not on same date, and 3) no influenza vaccine received within 1 week before to 1 week after receipt of the XBB.1.5-containing vaccine. Variant-specific comparative effectiveness was assessed at 6 weeks of follow-up and by stratifying calendar time according to before (XBB-lineage, mainly EG.5.1, prevailing) and after (BA.2.86-lineage, mainly JN.1, prevailing) 30 November 2023. We were only able to follow-up for six weeks for the variant-specific analysis given the short overlap in time where the XBB-lineage predominated and the vaccine was administered. Changes in the comparative vaccine effectiveness during follow-up (i.e., waning vaccine immunity) were assessed by stratifying follow-up in three-week intervals and subsequently fitting a linear regression, where the slope coefficient represented the per three-week percentage point change in comparative vaccine effectiveness.[12] Lastly, we carried out a sensitivity analysis starting follow-up three weeks after the index date to further reduce the potential of transient healthy vaccinee effect around the time of vaccination as well as possible spill-over effect from a delay between infection and onset of severe disease.

### Patient and public involvement

No patients or members of the public were formally involved in defining the research question, study design, or outcome measures, or in the conduct of the study owing to privacy constrains, funding restrictions, and the short timeline during which the study was conducted.

## RESULTS

### Study populations

Table 1 shows cohort characteristics before and after matching; supplementary figure S2 shows distributions of age and index date in density plots. Prior to matching, the source population consisted of 3,891,978 individuals eligible for vaccination with XBB.1.5-containing covid-19 mRNA vaccine as of study start, 1 October 2023. A total of 3,066,104 monovalent XBB.1.5-containing covid-19 mRNA vaccines were administered during the study period. The matched study cohorts comprised 1,867,448 recipients of an XBB.1.5-containing vaccine during the study period (mean age 75.4 years, standard deviation 7.4 years; 54.3% females; 554,638 from Denmark, 515,538 from Finland, and 797,272 from Sweden), matched with 1,867,448 non-recipients. Most XBB.1.5-containing vaccines were administered as a fifth covid-19 vaccine dose (53.2%) and during October 2023 in Denmark and November 2023 in Finland and Sweden. The distribution of the matched cohort characteristics was similar to the distribution observed prior to matching.

**Table 1.** Cohort characteristics before and after matching of XBB.1.5-containing vaccine recipients and non-recipients aged ≥65 years in Denmark, Finland, and Sweden, 1 October 2023 to 29 February 2024.^a^.

|  | Before matching |  | After matching |  |
| --- | --- | --- | --- | --- |
|  | XBB.1.5-containing vaccine recipients | XBB.1.5-containing vaccine non-recipients<sup>b</sup> | XBB.1.5-containing vaccine recipients | XBB.1.5-containing vaccine non-recipients |
| Total individuals | 3,066,104 | 3,891,978 | 1,867,448 | 1,867,448 |
| Denmark | 928,226 | 1,066,797 | 554,638 | 554,638 |
| Finland | 718,164 | 1,079,425 | 515,538 | 515,538 |
| Sweden | 1,419,714 | 1,745,756 | 797,272 | 797,272 |
| Mean age (SD), years | 75.9 (7.3) | 75.5 (7.6) | 75.4 (7.4) | 75.4 (7.4) |
| Female sex | 1,663,305 (54.2) | 2,112,686 (54.3) | 1,014,154 (54.3) | 1,014,154 (54.3) |
| Dose at which the XBB.1.5-containing vaccine was received |  |  |  |  |
| Fifth dose | 1,426,692 (46.5) | NA | 992,701 (53.2) | NA |
| Sixth dose | 1,247,379 (40.7) | NA | 699,321 (37.4) | NA |
| Seventh dose | 389,867 (12.7) | NA | 174,593 (9.3) | NA |
| Eighth dose | 2,166 (0.1) | NA | 833 (0.0) | NA |
| Severe covid-19 risk group | 656,591 (21.4) | 913,260 (23.5) | 436,938 (23.4) | 436,938 (23.4) |
| Healthcare workers | 131,005 (4.3) | 174,896 (4.5) | 82,375 (4.4) | 82,375 (4.4) |
| Autoimmune related condition | 147,816 (4.8) | 179,573 (4.6) | 83,792 (4.5) | 82,642 (4.4) |
| Cancer | 251,831 (8.2) | 311,467 (8.0) | 145,441 (7.8) | 143,913 (7.7) |
| Chronic pulmonary disease | 134,147 (4.4) | 167,056 (4.3) | 79,874 (4.3) | 77,840 (4.2) |
| Cardiovascular condition | 357,062 (11.6) | 443,241 (11.4) | 208,005 (11.1) | 207,828 (11.1) |
| Diabetes | 258,747 (8.4) | 339,724 (8.7) | 159,905 (8.6) | 162,666 (8.7) |
| Renal disease | 86,194 (2.8) | 112,311 (2.9) | 48,296 (2.6) | 50,974 (2.7) |
| Number of comorbidities |  |  |  |  |
| 0 | 2,616,364 (85.3) | 3,334,013 (85.7) | 1,608,115 (86.1) | 1,608,115 (86.1) |
| 1 | 415,034 (13.5) | 514,366 (13.2) | 241,565 (12.9) | 241,565 (12.9) |
| 2 | 33,311 (1.1) | 41,812 (1.1) | 17,340 (0.9) | 17,340 (0.9) |
| $\geq 3$ | 1,395 (0.0) | 1,787 (0.0) | 428 (0.0) | 428 (0.0) |

NA denotes not applicable SD standard deviation. <sup>a</sup>Values are numbers (percentages) unless stated otherwise. <sup>b</sup>Individuals eligible for XBB.1.5-containing vaccine vaccination as of study start, 1 October 2023.

### Effectiveness of XBB.1.5-containing vaccines

Figure 1 shows the 12-week cumulative incidences of hospital admission and death related to covid-19 in XBB.1.5-containing vaccine recipient versus matched non-recipient individuals from 1 week after the vaccination date. Overall, cumulative incidences of severe covid-19 outcomes were low for both recipients and non-recipients. The risk of admission to hospital with covid-19, however, was lower for individuals who had received an XBB.1.5-containing vaccine compared with individuals who had not (930 vs 2,551 events) corresponding to an estimated comparative vaccine effectiveness of 60.6% (95% confidence interval, 46.1% to 75.1%) and risk difference per 100,000 individuals of –191.1 (95% confidence interval, –332.1 to –50.2) at week 12 (Table 2). For covid-19 related death (301 vs 1,326), the comparative vaccine effectiveness was 77.9% (69.2% to 86.7%) and the risk difference per 100 000 individuals was –109.2 (–118.1 to –100.2). While the comparative vaccine effectiveness was similar irrespective of sex, age group, and number of previous covid-19 vaccine doses the absolute risk difference point estimate was larger in males, in individuals aged ≥75 years, and in individuals with higher number of covid-19 vaccine doses previously received, but the 95% confidence intervals largely overlapped for covid-19 hospital admission (e.g., risk differences against admission to hospital with covid-19 were –194.6 [–271.6 to –117.6] and –86.4 [–160.3 to –12.5] per 100,000 individuals aged ≥/<75 years, respectively). No apparent differences in the comparative effectiveness were observed according to seasonal influenza vaccine co-administration, but the 95% confidence intervals were more imprecise for this subgroup analysis. Variant-specific comparative effectiveness estimates against covid-19 related hospital admission and death at 6 weeks of follow-up were similar during XBB-lineage and BA.2-lineage period predominance; the comparative vaccine effectiveness point estimates were slightly higher against the XBB-lineage but the 95% confidence intervals overlapped.

**Figure 1.**
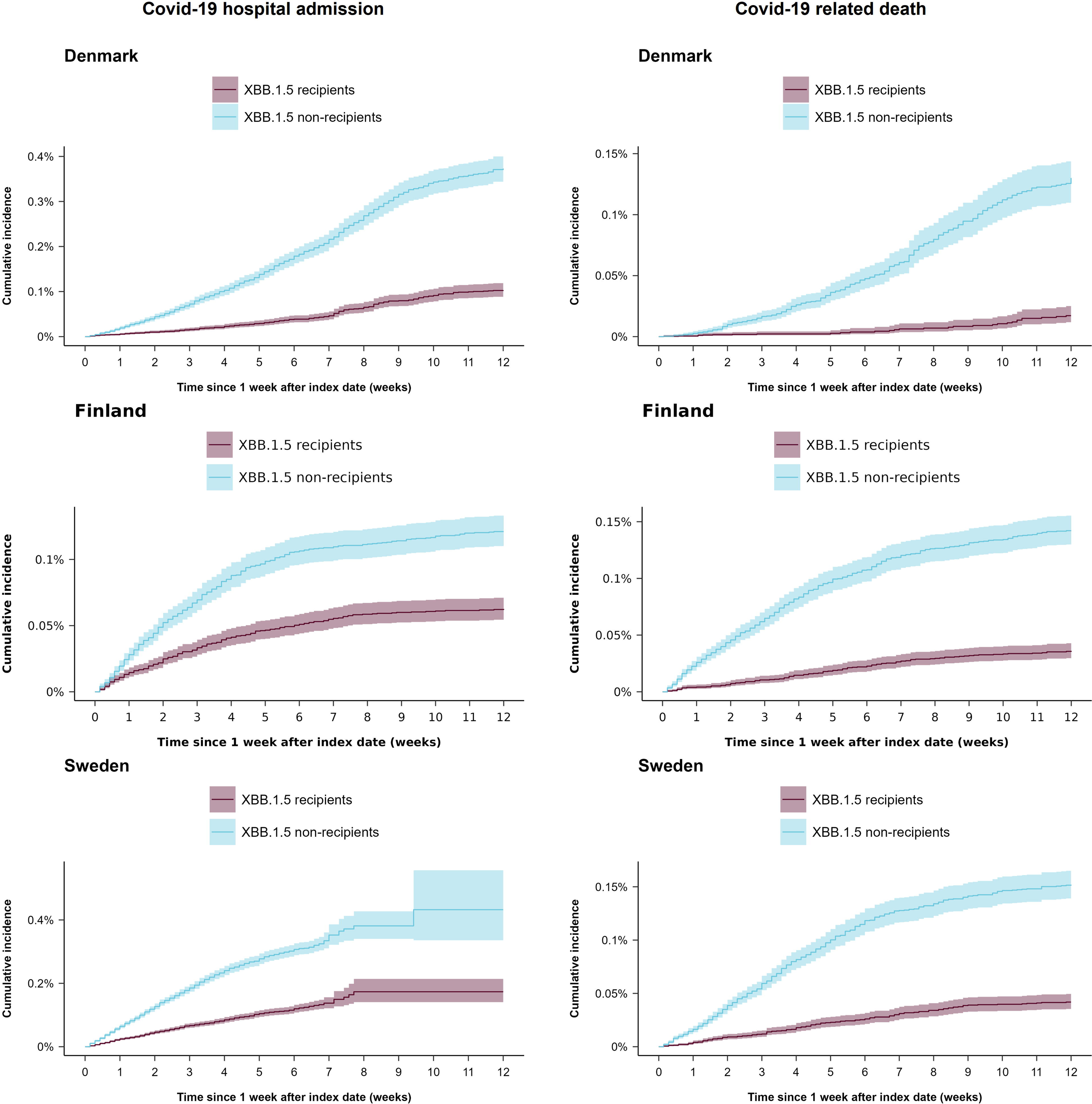
Cumulative incidence curves of admission to hospital and death related to covid-19, comparing recipients of a monovalent XBB.1.5-containing covid-19 mRNA vaccine during autumn and winter 2023-2024 with matched non-recipients. XBB.1.5 denotes XBB.1.5-containing vaccine. Matched XBB.1.5-containing vaccine recipients and non-recipients pairs were followed for a total of 12 weeks after immunisation (defined as 1 week after the day of vaccination).

**Table 2.** Risk of hospital admission and death related to covid-19 comparing XBB.1.5-containing vaccine recipients with non-recipients aged ≥65 years in Denmark, Finland, and Sweden, 1 October 2023 to 29 February 2024.^a^.

|  | Contributing countries | Events/person-years |  | Risk difference (95% CI) per 100,000 individuals | Comparative vaccine effectiveness (95% CI), % |
| --- | --- | --- | --- | --- | --- |
|  |  | XBB.1.5-containing vaccine recipients | XBB.1.5-containing vaccine non-recipients |  |  |
| Covid-19 hospital admission |  |  |  |  |  |
| All | DK, FI, SE | 930/170,115 | 2,551/168,911 | -191.1 (-332.1 to -50.2) | 60.6 (46.1 to 75.1) |
| Female | DK, FI, SE | 401/93,130 | 1,173/92,527 | -160.6 (-301.5 to -19.8) | 62.4 (47.9 to 77.0) |
| Male | DK, FI, SE | 529/76,985 | 1,378/76,385 | -214.3 (-366.1 to -62.5) | 58.8 (44.2 to 73.5) |
| Age <75 years | DK, FI, SE | 189/88,252 | 532/88,119 | -86.4 (-160.3 to -12.5) | 58.3 (42.1 to 74.6) |
| Age ≥75 years | DK, FI, SE | 741/81,863 | 2,019/80,792 | -194.6 (-271.6 to -117.6) | 62.0 (47.5 to 76.4) |
| XBB.1.5-containing vaccine received as fifth dose | DK, FI, SE | 388/108,684 | 1,197/107,995 | -156.2 (-293.6 to -18.8) | 64.6 (51.0 to 78.1) |
| XBB.1.5-containing vaccine received as sixth dose | DK, FI, SE | 385/53,312 | 965/52,914 | -188.4 (-361.6 to -15.2) | 57.0 (41.6 to 72.4) |
| XBB.1.5-containing vaccine received as seventh dose<sup>b</sup> | FI, SE | 156/8,086 | 387/7,969 | -248.9 (-490.4 to -7.4) | 44.4 (20.2 to 68.7) |
| Influenza vaccine received on same day | DK, FI | 376/108,174 | 1,100/107,323 | -169.1 (-416.4 to 78.3) | 61.5 (38.6 to 84.4) |
| Influenza vaccine received within 1 week | DK, FI | 13/3,040 | 38/3,014 | -148.6 (-417.1 to 119.8) | 54.3 (20.0 to 88.7) |
| No concurrent influenza vaccine received | DK, FI | 42/13,295 | 91/13,229 | -122.8 (-372.1 to 126.5) | 58.5 (31.6 to 85.4) |
| XBB-sublineages prevailing<sup>c</sup> | DK, FI, SE | 396/61,813 | 1,290/61,646 | -154.3 (-313.9 to 5.4) | 73.6 (60.4 to 86.7) |
| BA.2.86-sublineages prevailing<sup>c</sup> | DK, FI | 147/34,112 | 318/33,949 | -158.2 (-318.1 to 1.8) | 56.6 (42.8 to 70.4) |
| Covid-19 death |  |  |  |  |  |
| All | DK, FI, SE | 301/203,402 | 1,326/201,981 | -109.2 (-118.1 to -100.2) | 77.9 (69.2 to 86.7) |
| Subgroups |  |  |  |  |  |
| Female | DK, FI, SE | 146/110,653 | 596/109,938 | -87.3 (-98.3 to -76.2) | 76.9 (67.6 to 86.2) |
| Male | DK, FI, SE | 155/92,749 | 730/92,043 | -133.3 (-147.7 to -118.8) | 78.9 (69.9 to 87.9) |
| Age <75 years | DK, FI, SE | 35/105,912 | 159/105,780 | -25.9 (-32.1 to -19.8) | 77.5 (65.6 to 89.5) |
| Age ≥75 years | DK, FI, SE | 266/97,490 | 1,167/96,201 | -201.6 (-219.4 to -183.9) | 78.0 (69.3 to 86.8) |
| XBB.1.5-containing vaccine received as fifth dose | DK, FI, SE | 113/124,819 | 533/124,148 | -75.9 (-111.8 to -40.1) | 77.7 (67.5 to 87.9) |
| XBB.1.5-containing vaccine received as sixth dose | DK, FI, SE | 140/67,095 | 563/66,576 | -141.4 (-180.7 to -102.2) | 76.9 (66.4 to 87.4) |
| XBB.1.5-containing vaccine received as seventh dose<sup>b</sup> | SE | 48/11,230 | 225/11,000 | -322.4 (-396.6 to -248.1) | 82.1 (68.8 to 95.5) |
| Influenza vaccine received on same day | DK, FI | 133/108,526 | 670/107,752 | -112.9 (-125.2 to -100.5) | 80.8 (69.0 to 92.6) |
| Influenza vaccine received within 1 week | DK, FI | 6/3,044 | 14/3,022 | -75.5 (-146.1 to -4.8) | 73.5 (42.6 to 100.0) |
| No concurrent influenza vaccine received | DK, FI | 10/13,339 | 57/13,280 | -80.1 (-108.8 to -51.5) | 83.8 (67.7 to 99.9) |

**Table 2. Risk of hospital admission and death related to covid-19 comparing XBB.1.5-containing vaccine recipients with non-recipients aged $\geq 65$ years in Denmark, Finland, and Sweden, 1 October 2023 to 29 February 2024.<sup>a</sup>**
|  | Contributing countries | Events/person-years |  | Risk difference (95% CI) per 100,000 individuals | Comparative vaccine effectiveness (95% CI), % |
| --- | --- | --- | --- | --- | --- |
|  |  | XBB.1.5-containing vaccine recipients | XBB.1.5-containing vaccine non-recipients |  |  |
| XBB-sublineages prevailing<sup>c</sup> | DK, FI, SE | 70/61,934 | 444/61,796 | -45.7 (-54.3 to -37.1) | 87.5 (80.3 to 94.6) |
| BA.2.86-sublineages prevailing<sup>c</sup> | DK, FI, SE | 131/63,577 | 572/63,269 | -74.5 (-82.3 to -66.6) | 77.5 (71.4 to 83.6) |

CI denotes confidence interval, DK Denmark, FI Finland, and SE Sweden. <sup>a</sup>Individuals were followed for 12 weeks (from 1 week after the vaccination date), except for the estimates by prevailing omicron sublineages where individuals were followed for 6 weeks. <sup>b</sup>Risk of the covid-19 outcomes could not be separately studied in subgroups of individuals where the XBB.1.5-containing vaccine was received as an eighth dose due to too few events. <sup>c</sup>Assessed at 6 weeks after start of follow-up; see supplementary table S6 for overall 6-week of follow-up results.

Figure 2 shows the comparative vaccine effectiveness stratified by 3-week intervals and the trend line, representing the per 3-three week change in effectiveness during follow-up. Estimates suggested slightly higher initial protection, with comparative vaccine effectiveness of 65.2% (50.6% to 79.6%) against covid-19 related hospital admission and 82.7% (79.2% to 86.2%) against covid-19 related death at 3 weeks of follow-up with subsequent gradual waning of –2.0 (95% confidence interval, –8.8 to 4.8) and –3.7 (–7.5 to 0.2) percentage points every 3 weeks, respectively.

**Figure 2.**
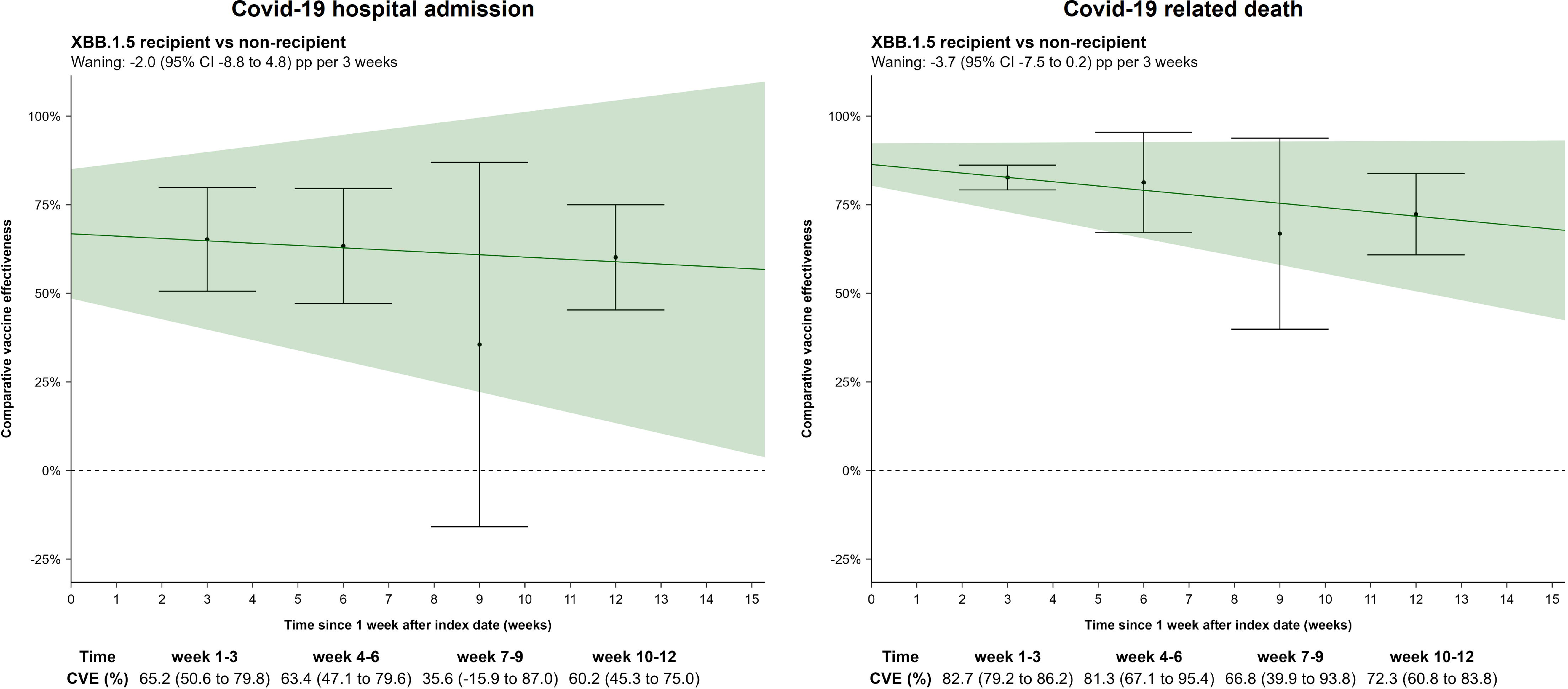
Waning comparative vaccine effectiveness against admission to hospital and death related to covid-19, comparing recipients of a monovalent XBB.1.5-containing covid-19 mRNA vaccine during autumn and winter. 2023**-**2024 **with matched non-recipients, stratifying follow-up in 3-week intervals.** CVE denotes comparative vaccine effectiveness. Waning estimates represent the trend line in the per 3-week comparative vaccine effectiveness estimates.

Starting follow-up 3 weeks after vaccination did not change the overall findings (supplementary table S5).

## DISCUSSION

In this multicohort analysis across three Nordic countries of individuals aged ≥65 years, we observed lower rates of hospital admission and death related to covid-19 associated with receipt of a monovalent XBB.1.5-containing covid-19 mRNA vaccine dose compared with no receipt during autumn and winter 2023-2024. Specifically, we estimated a comparative vaccine effectiveness of 60.6% against covid-19 related hospital admission and of 77.9% against death at 12 weeks follow-up. In addition, we found that the comparative vaccine effectiveness of the XBB.1.5-containing vaccine did not differ between sex, age groups, number of previous covid-19 vaccine doses, and if seasonal influenza vaccination was co-administered, nor between periods of XBB-lineage (EG.5.1) and BA.2.86-lineage (JN.1) predominance. Moreover, we observed that the protection afforded waned only modestly during follow-up with a well-preserved comparative vaccine effectiveness at end of 12 weeks.

### Comparison with other studies

Our results support that variant-updated covid-19 vaccination with the XBB.1.5-containing vaccine targeting the elderly, successfully prevented a significant number of severe covid-19 events across three Nordic countries during autumn and winter 2023-2024. Our findings also align well with the available early short-term vaccine effectiveness estimates of the XBB.1.5-containing vaccine.[5–8] With 2.5-week data from 8 to 26 October 2023 and an average follow-up of 9.9 days, a cohort analysis in Denmark found an early high short-term vaccine effectiveness of 76% against covid-19 related hospitalisation associated with the XBB.1.5-containing covid-19 vaccine.[6] Similar early-season vaccine effectiveness was recently reported by the VEBIS (Vaccine Effectiveness Burden and Impact Studies) project of >66% against covid-19 related hospitalisation and death (data until 25 November 2023) and from the Netherlands of 71% against covid-19 related hospitalisation (data until 5 December 2023).[7,8] Using a test-negative case-control design, reports from the UK found that the XBB.1.5-containing vaccine was associated with a comparative vaccine effectiveness peak of 55% against covid-19 related hospitalisation 2 to 4 weeks after vaccination in individuals aged ≥65 years.[5,13] Our findings also suggest that the protection afforded by the XBB.1.5-containing vaccine was initially higher—with an estimated overall comparative vaccine effectiveness at 3 weeks of follow-up (that is, 4 weeks from the vaccination date) of 65% and 82% against covid-19 related hospital admission and death, respectively.

With data up until 29 February 2024, our primary analysis provides an evaluation of the comparative effectiveness of the entire autumn and winter 2023-24-season. We further add to the current evidence by showing that the initial peak in protection was well-preserved at 12 weeks, suggestive of only modest waning of effect and by analysing a range of subgroups. Additionally, we show that the comparative effectiveness of the XBB.1.5-containing vaccine against severe covid-19 outcomes was relatively similar between the periods of XBB- and BA.2.86-sublineages predominance. Estimates from the UK suggested that vaccination with either the XBB.1.5-containing or the bivalent BA.4-5 booster mRNA covid-19 vaccine (i.e., XBB.1.5-containing vaccine was not separately studied) had a higher relative protection against hospitalisation with XBB-sublineages.[13] Our results similarly tended toward slightly higher comparative vaccine effectiveness during the XBB-rather than the BA.2.86-sublineage predominance period, but the 95% confidence intervals largely overlapped and this potential difference was also not reflected in the absolute risk estimates. It should be noted, however, that any indirect comparison of the comparative effectiveness of a covid-19 vaccine against different SARS-CoV-2 strains is inherently affected by the strong correlation with calendar time as well as changes in background population transmission rates.

In contrast to the abovementioned studies, we present estimates of benefits of vaccination in absolute terms. Specifically, the comparative effectiveness estimates of our primary analysis corresponds to 191.1 (95% confidence interval, 50.2 to 332.1) hospital admissions and 109.2 (100.2 to 118.1) deaths related to covid-19 prevented per 100,000 individuals vaccinated with an XBB.1.5-containing vaccine within our Nordic population. Moreover, while the relative comparative vaccine effectiveness measures were alike, we show that the absolute benefit from vaccination with the XBB.1.5-containing vaccine varies across subgroups: being higher among males, those aged ≥75 years, and those having received more prior covid-19 vaccine doses—reflecting higher background risk for these subpopulations. Consequently, accompanying the relative with absolute measures provides health authorities, clinicians, and patients with a more complete evaluation of the benefits of vaccination.

### Strengths and limitations of study

Our study has limitations. First, our outcome ascertainment likely also captured a proportion of cases in which the infection with SARS-CoV-2 only partly contributed to or coincided with the timing of admission to hospital or death. Second, as part of our outcome definitions, individuals were required to have a positive PCR test for SARS-CoV-2, and therefore, individuals who were hospitalised or who died because of covid-19 but were not tested were missed. Third, we cannot exclude the possibility of residual confounding. To bias our results, unmeasured confounding factors would need to be unevenly distributed between compared groups and not indirectly taken into account by the set of included covariates (that is, proxies). In addition, by study design, the matched cohort also included pairs where the non-recipient reference individual received an XBB.1.5-containing vaccine later than the assigned index date, and opting for earlier relative to later in-season vaccination could represent differences in severe covid-19 risk. We attempted to mitigate these concerns by using an active comparison group and exact matching on potential key confounders. If not addressed adequately in our design, these biases would most likely tend to skew our results toward the null. Fourth, the majority of individuals who received an XBB.1.5-containing vaccine also received their seasonal influenza vaccine on the same day. Hence, the 95% confidence interval for the other seasonal influenza vaccination status subgroups were more imprecise. In addition, this also means that our main estimates primarily reflect the co-administration with the seasonal influenza vaccine. This is reassuring, since co-administration has been speculated to blunt the immune response,[14–19] and if that is the case, our estimates are likely conservative.

As we studied the general population of adults aged ≥65 years, our results should be highly generalizable to other similar populations targeted for vaccination with the XBB.1.5-containing vaccine during autumn and winter 2023-2024. Accordingly, these results may only indirectly support evaluations within populations not herein studied. Similarly, analyses were carried out for the autumn and winter 2023-2024 season during which the XBB-sublineages (particularly EG.5.1; until around end November 2023) and subsequently the BA.2.86-sublineages (particularly JN.1) were prevailing. Consequently, the transportability of our results to protection potentially conferred against severe covid-19 caused by other SARS-CoV-2 subvariants is unknown.

## CONCLUSIONS

We found that the monovalent XBB.1.5-containing covid-19 mRNA vaccine reduced the rates of hospital admission and death related to covid-19 among individuals aged ≥65 years during autumn and winter 2023-2024 across the three Nordic countries of Denmark, Finland, and Sweden. We observed that the protection afforded did not differ between sex, age, number of previous covid-19 vaccine doses, and seasonal influenza vaccination co-administration subgroups, nor between periods of XBB-lineage (EG.5.1) and BA.2.86-lineage (JN.1) predominance. While the protection was highest during the first weeks after vaccination, it was well-preserved at end of the 12 weeks of follow-up.

## Data Availability

Owing to data privacy regulations in each country, the raw data cannot be shared.

## ACKNOWLEDGMENT SECTION

### Contributors

NWA, EMT, and AH conceptualized the study. NWA drafted the manuscript. EMT, NP, and JP did the statistical analysis. All authors interpreted the results and critically reviewed the manuscript. AH supervised the study. The corresponding author attests that all listed authors meet authorship criteria and that no others meeting the criteria have been omitted. NA, EMT, and AH are the guarantors.

### Funding

This research was supported by the European Medicines Agency. The funders had no role in deciding the study design; in the collection, analysis, and interpretation of data; in the writing of the report; or in the decision to submit the article for publication. This document expresses the opinion of the authors of the paper and may not be understood or quoted as being made on behalf of, or reflecting the position of, the European Medicines Agency or one of its committees or working parties.

### Ethics

The Danish analyses were performed as surveillance activities analyses as part of the advisory tasks of the governmental institution Statens Serum Institut (SSI) for the Danish Ministry of Health. SSI’s purpose is to monitor and fight the spread of disease in accordance with section 222 of the Danish Health Act. According to Danish law, national surveillance activities conducted by SSI do not require approval from an ethics committee. Both the Danish Governmental law firm and the compliance department of SSI have approved that the study is fully compliant with all legal, ethical, and IT-security requirements and there are no further approval procedures required for such studies.

For the Finnish analyses, by Finnish law, the Finnish Institute for Health and Welfare (THL) is the national expert institution to carry out surveillance of the impact of vaccinations in Finland (Communicable Diseases Act, https://www.finlex.fi/en/laki/kaannokset/2016/en20161227.pdf). Neither specific ethical approval of this study nor informed consent from the participants were needed.

The Swedish analyses were conducted under the Swedish Ethical Review Authority approval 2020-06859, 2021-02186 and conformed to the principles embodied in the Declaration of Helsinki. Register-based studies (like this) in Sweden are exempt from obtaining consent to participate.

### Data sharing

Owing to data privacy regulations in each country, the raw data cannot be shared. **Transparency:** The lead author (the manuscript’s guarantor) affirms that this manuscript is an honest, accurate, and transparent account of the study being reported; that no important aspects of the study have been omitted; and that any discrepancies from the study as planned (and, if relevant, registered) have been explained.

### Dissemination to participants and related patient and public communities

Studied participants were anonymised in the data sources used; therefore, direct dissemination to study participants is not possible. The study results will be disseminated to the public and health professionals by a press release written using layman’s terms.

