## supplementary tables S1-S2 for "Comparative effectiveness of the monovalent XBB.1.5-containing covid-19 mRNA vaccine across three Nordic countries"

### Supplementary Text. Ethics approval/exempt within each country.

**Denmark:** The Danish analyses were performed as surveillance activities analyses as part of the advisory tasks of the governmental institution Statens Serum Institut (SSI) for the Danish Ministry of Health. SSI's purpose is to monitor and fight the spread of disease in accordance with section 222 of the Danish Health Act. According to Danish law, national surveillance activities conducted by SSI do not require approval from an ethics committee. Both the Danish Governmental law firm and the compliance department of SSI have approved that the study is fully compliant with all legal, ethical, and IT-security requirements and there are no further approval procedures required for such studies.

**Finland:** By Finnish law, the Finnish Institute for Health and Welfare (THL) is the national expert institution to carry out surveillance of the impact of vaccinations in Finland (Communicable Diseases Act, https://www.finlex.fi/en/laki/kaannokset/2016/en20161227.pdf). Neither specific ethical approval of this study nor informed consent from the participants were needed.

**Sweden:** The Swedish analyses were conducted under the Swedish Ethical Review Authority approval 2020-06859, 2021-02186 and conformed to the principles embodied in the Declaration of Helsinki. Register-based studies (like this) in Sweden are exempt from obtaining consent to participate.

**Supplementary Table S1. Overview of utilised registers.**

| Country/data source | Details |
|---|---|
| **Denmark** | |
| The Civil Registration System[1] | The register provides the unique personal identifier for all permanent residents of Denmark that allows linkage between all Danish health care registers and civil registrations systems. In addition, it holds general demographic information such as birthdate and sex as well as continuously updated information and dates on historical addresses, immigration and emigration status, and death. |
| The Danish Vaccination Register[2] | The register holds information on all vaccinations administered in Denmark including vaccination date, type/trade name, dose, and product batch number ever since Nov 15, 2015 (where reporting to the register became mandatory). Specifically related to this study, the Danish Health Agency have provided the governmentally assigned Covid-19 vaccine priority groups that were prioritized groups according to the risk of severe infection as well as whether being health and social care workers. |
| The Danish Microbiology Database[3] | Information on positive PCR tests for SARS-CoV-2 are obtainable via The Danish Microbiology Database (MiBa) that has data on all microbiology samples analysed at Danish microbiology departments as well as test results, date of sampling, date of analysis, type of test, and interpretation of test. The SARS-CoV-2 PCR tests are freely available to all individuals in Denmark regardless of symptoms status. |
| The National Patient Register[4] | The register holds information on all hospital contacts in Denmark including the duration of the contact, and diagnoses, which are assigned by the treating physician and registered according to ICD-10 classification system (since 1994). |
| **Finland** | |
| Finnish Population Information System[5] | The register is held by the Digital and Population Data Services Agency and contains personal data on all permanent residents in Finland such as the unique personal identifier, date of birth, place of residence, date of death, and date of immigration, and emigration. |
| Register of Social Assistance[6] | The register is held by the Finnish Institute for Health and Welfare and contains information on individuals receiving long-term care and/or social assistance (in e.g., nursing homes, people's own homes or other institutions) including social rehabilitation. |
| Social and Healthcare Professionals Register[7] | The register holds data on individuals right to act as health care personnel. |
| National Vaccination Register[8] | The register is based on the Register of Primary Health Care Visits and contains information on all Covid-19 vaccinations administered in Finland including date of vaccination, batch number, and trade name. |
| National Infectious Diseases Register[9] | The register is held by the Finnish Institute for Health and Welfare and contains information on notifiable diseases in accordance with the Finnish Communicable Diseases Act that must be reported by the laboratories and the treating-physicians, or the physician performing an autopsy and hold information on sample dates of all laboratory-confirmed SARS-CoV-2 infections in Finland |
| National Care Register for Health Care[10] | The register is held by the Finnish Institute for Health and Welfare and comprises information on all inpatient and outpatient hospital contacts in Finland, including admission and discharge dates, whether hospitalization was planned or acute, codes for discharge diagnoses (according to ICD-10) and surgical procedures, and whether discharged as deceased, to own private residence or other health care facilities. |
| Special Reimbursement Register and Prescription Centre database | These databases are maintained by the Finnish Social Insurance Institution. The Special Reimbursement Register holds information on individuals entitled to special reimbursement for medical expenses. The Prescription Centre database holds information on individuals using selected medications of interest. |

| Country/data source | Details |
|---|---|
| Register of Primary Health Care Visits[11] | The register is held by Finnish Institute for Health and Welfare and holds data on all primary health care services delivered in Finland. |
| **Sweden** | |
| The Total Population Register[18] | The register is held by Statistics Sweden and contains data on the unique personal identifier assigned to all individuals in Sweden plus general demographic information such as date of birth, sex, country of birth, place of residence, and date of immigration and emigration. |
| The Cause of Death Register[19] | The register holds information on date of death and underlying as well as contributing causes of death. |
| The Longitudinal Integrated Database For Health Insurance And Labour Market Studies (LISA)[20] | The database is held by Statistics Sweden and holds many socioeconomic variables such as data on occupation which we used to identify whether individuals were health care personnel. |
| Register On Persons In Nursing Homes[21] | The register is held by the National Board of Health and Welfare and holds data on nursing care given in either nursing homes, own homes or other institutions to elderly and/or persons with physical, psychiatric or intellectual disabilities. |
| The National Vaccination Register[22] | The register is held by the Public Health Agency of Sweden and contains information on administered Covid-19 vaccines in Sweden including data on date of administration, the specific vaccine products, substance, formulation, batch number and dose number (for repeated doses). |
| Register On Surveillance Of Notifiable Communicable Diseases (Sminet)[23] | The register is held by the Public Health Agency of Sweden and contains information on notifiable diseases (for which reporting is mandatory) reported by either the analysis performing laboratories, the treating physician or autopsy performing physician, in accordance with the Swedish Communicable Diseases Act. Data included are e.g., date of disease occurrence, date of testing, date of positive test, and diagnoses. |
| The Swedish Patient Register[24,25] | The register is held by the National Board of Health and Welfare and comprises data on all in- and outpatient hospital specialist care in Sweden including data on dates of admission and discharge, whether hospitalization was planned or acute, codes for discharge diagnoses (recorded according to ICD-10-SE) and surgical procedures, whether discharged as deceased, to own private residence or other health care facilities, and type of department. |

The nationwide registries were linked via the unique personal identifier that is assigned to all residents within the respective Nordic country at either birth or immigration. Consequently, all utilised data were collected on the individual level. The healthcare systems in the Nordic countries are universal and tax-financed meaning the healthcare services are either freely available to all or subsidised so that all individuals pay only a fixed-based minimum irrespective of the actual services provided and costs. We had full data availability for all variables during the study period and as reporting to national registers is mandatory/structurally implemented, this provides a near-complete follow-up of all residents over time.

**Supplementary Table S2. Covariate definitions.**

| Variable | Country | Data source and details | Values/codes |
|---|---|---|---|
| Age | Denmark | *The Civil Registration System.* Recorded birth year. Age defined as the study start date (1 October 2023) minus birth year. | Categorical (for adjustment, using birth year): 5-year bins<br>Binary (for stratification): 65-74/≥ 75 years |
| | Finland | *The Finnish Population Information System.* Recorded birth year. Age defined as the study start date minus birth year. | |
| | Sweden | *The Total Population Register.* Recorded birth year. Age defined as the year of study start (2023) minus birth year. | |
| Sex | Denmark | *The Civil Registration System.* Defined as registered sex. | Binary: male, female |
| | Finland | *The Finnish Population Information System.* Defined as registered sex. | |
| | Sweden | *The Total Population Register.* Defined as registered sex. | |
| Calendar time period of last mutual vaccine dose | Denmark | *The Danish Vaccination Register.* Defined by the date where the respective vaccine dose examined was administered (e.g., fourth or fifth dose). | Categorical: monthly-bins from 27 December 2020-31 January 2021 to February 2024 |
| | Finland | *The National Vaccination Register.* Defined by the date where the respective vaccine dose examined was administered (e.g., fourth or fifth dose). | |
| | Sweden | *The National Vaccination Register.* Defined by the date where the respective vaccine dose examined was administered (e.g., fourth or fifth dose). | |
| Region of residency | Denmark | *The Civil Registration System.* Defined by last known address at the study start date. | Categorical: Denmark, 5 levels; Finland, 5 levels; Sweden, 9 levels |
| | Finland | *The Finnish Population Information System.* Defined by last known municipality of residence at the study start date. | |
| | Sweden | *The Total Population Register.* Defined by last known address at the study start date. | |
| Covid-19 risk groups | Denmark | *The Danish Vaccination Register.* Defined as governmentally assigned covid-19 vaccine priority groups, prioritized according to the risk of severe covid-19 as well as whether being health and social care workers (last update 24 May 2021). | Categorical (3 levels): Severe covid-19 risk group, healthcare personnel, others |
| | Finland | *Register of Social Assistance.* Severe covid-19 risk group was defined as vulnerable individuals in 24-hours care (binary status per 27 December 2021).<br><br>*Care register for Health Care (data since 1.1.2015), Special Reimbursement Register (data from 1.1.2018 to 27.12.2020) Prescription Centre database (data from 1.1.2018 to 27.12.2020.* Covid-19 risk group was defined on the basis of national vaccination recommendation [9]. | |
| | Sweden | *Register on persons in nursing homes.* Severe covid-19 risk group was defined as vulnerable individuals being residents at nursing homes (binary status as of 31 December 2020) | |

| Variable | Country | Data source and details | Values/codes |
|---|---|---|---|
| | | *The Longitudinal integrated database for health insurance and labour market studies.* Healthcare personnel defined as healthcare worker occupation status as of 31 October 2018 (binary). | |
| Chronic pulmonary disease | Denmark | *The National Patient Register.* Defined as primary diagnoses regardless of type of hospital contact registered prior to start of the study period (look-back 3 years). | Binary: yes/no (ICD-10 codes: J40-J47, J60–J67, J684, J701, J703, J841, J920, J961, J982, J983) |
| | Finland | *Care register for Health Care.* Defined as primary or secondary diagnoses registered prior to the start of the study period. | Binary: yes/no (ICD-10 codes: J41-J44, J47) |
| | Sweden | *National Patient Register.* Defined as any recorded ICD-10 diagnosis during inpatient or outpatient contact and prior to the start of the study period (look-back 3 years). | Binary: yes/no (ICD-10 codes: E84, J41-J47, J84, J98) |
| Cardiovascular conditions | Denmark | *The National Patient Register.* Defined as primary diagnoses regardless of type of hospital contact registered prior to the start of the study period (look-back 3 years). | Binary: yes/no (ICD-10 codes: I110, I130, I132, I20-I23, I420, I426-I429, I48, I500-I503, I508, I509) |
| | Finland | *Care register for Health Care, Register of Primary Health Care Visits, Special Reimbursement Register and Prescription Centre database.* Defined as primary or secondary diagnoses prior to the start of the study period. | Binary: yes/no (ICD-10 codes: I11–I13, I15, I20–I25) |
| | Sweden | *National Patient Register.* Defined as any recorded ICD-10 diagnosis during inpatient or outpatient contact and prior to the start of the study period (look-back 3 years). | Binary: yes/no (ICD-10 codes: I05-I09, I110, I20-I28, I34-I37, I39, I42, I43, I46, I48-I50) |
| Diabetes | Denmark | *The National Patient Register.* Defined as primary diagnoses regardless of type of hospital contact registered prior to the start of the study period (look-back 3 years). | Binary: yes/no (ICD-10 codes: E10-E11) |
| | Finland | *Care register for Health Care, Register of Primary Health Care Visits, Special Reimbursement Register and Prescription Centre database.* Defined as primary or secondary diagnoses prior to the start of the study period or drug prescriptions before 27 December 2020. | Binary: yes/no (ICD-10 codes: E10, E11, E13-E14; ICPC-2 codes: T89, T90; ATC codes: A10A, A10B) |
| | Sweden | *National Patient Register and Swedish Prescribed Drug Register.* Defined as any recorded ICD-10 diagnosis during inpatient or outpatient contact and prior to the start of the study period or antidiabetic drugs use defined as ≥2 filled prescriptions (look-back 3 years). | Binary: yes/no (ICD-10 codes: E10-E14; ATC code: A10) |
| Autoimmunity-related conditions[a] | Denmark | *The National Patient Register.* Defined as primary diagnoses regardless of type of hospital contact registered prior to the start of the study period (look-back 3 years). | Binary: yes/no (ICD-10 codes: D510, D590, D591, D690, D693, D86, E050, E063, E271, E272, G122G, G35, G610, G700, I00, I01, K50, K51, K743, K900, L12, L40, L52, L80, L93, M05, M06, M08, M300, M313, M315, M316, M32, M33, M34, M35, M45) |
| | Finland | *Care register for Health Care, Special Reimbursement Register and Prescription Centre database.* Defined as primary or secondary diagnoses prior to the start of the | Binary: yes/no (ICD-10 codes: D7081, D7089, D80–D84, E250, E271, E272, E274, E310, E896, |

| Variable | Country | Data source and details | Values/codes |
|---|---|---|---|
| | | study period or drug prescriptions before 27 December 2020. | D86, K50, K51, L40, M02, M05–M07, M139, M45, M460, M461, M469, M941; ATC-codes: H02AB02, H02AB04, H02AB06, H02AB07, L01BA01, L01XC02, L04AA06, L04AA10, L04AA13, L04AA18, L04AA24, L04AA26, L04AA29, L04AA33, L04AA37, L04AB, L04AC, L04AD01, L04AD02, L04AX01, L04AX03) |
| | Sweden | *National Patient Register.* Defined as any recorded ICD-10 diagnosis during inpatient or outpatient contact and prior to the start of the study period (look-back 3 years). | Binary: yes/no (ICD-10 codes: D86, G35, K50, K51, L40, M05-M09, M13, M14, M45) |
| Cancer | Denmark | *The National Patient Register.* Defined as primary diagnoses regardless of type of hospital contact registered prior to the start of the study period (look-back 3 years). | Binary: yes/no (ICD-10 codes: C00–C85 (without C44), C88, C90-C96) |
| | Finland | *Care register for Health Care and Special Reimbursement Register.* Defined as primary or secondary diagnoses registered prior to the start of the study period (look-back 2 years) | Binary: yes/no (ICD-10 codes: C00–C43, C45–C80, C97, D05.1, D39) |
| | Sweden | *National Patient Register.* Defined as any recorded ICD-10 diagnosis during inpatient or outpatient contact and prior to the start of the study period (look-back 3 years). | Binary: yes/no (ICD-10 codes: C00-C96 (without C44), D45-D47) |
| Moderate to severe renal disease | Denmark | *The National Patient Register.* Defined as primary diagnoses regardless of type of hospital contact registered prior to the start of the study period (look-back 3 years). | Binary: yes/no (ICD-10 codes: I12, I13, N00–N05, N07, N11, N14, N17–N19, Q61) |
| | Finland | *Care register for Health Care.* Defined as primary or secondary diagnoses prior to the start of the study period. | Binary: yes/no (ICD-10 codes: I12, I13, N00–N05, N07, N08, N11, N14, N18, N19, E102, E112, E142) |
| | Sweden | *National Patient Register.* Defined as any recorded ICD-10 diagnosis during inpatient or outpatient contact and prior to the start of the study period (look-back 3 years). | Binary: yes/no (ICD-10 codes: I12, I13, N00-N05, N07, N11, N14, N17-N19, Q61) |
| 2023-2024 seasonal influenza vaccination | Denmark | *The Danish Vaccination Register.* Defined according to the date of influenza vaccine and XBB.1.5-containing vaccine vaccinations. | Categorical (for subgroup analysis only): co-administered on the same date; received influenza vaccine within 1 week before to 1 week after XBB.1.5-containing vaccine dose administration but no on same date; no influenza vaccine administered within 1 weeks before to 1 week after of XBB.1.5-containing vaccine dose administration. |
| | Finland | *The National Vaccination Register.* Defined according to the date of influenza vaccine (VaxigripTetra or InfluvacTetra) and XBB.1.5-containing vaccine vaccinations. | |
| | Sweden | Not available in Sweden. | |

[a]Autoimmunity-related conditions includes a range disorders such as inflammatory bowel diseases, diseases involving the blood, immune mechanism or endocrine systems, inflammatory rheumatic diseases, psoriasis, lupus erythematosus, multiple sclerosis; subject to country-specific definitions. The selected diagnosis codes to define comorbidities were country-specific, based on inputs from national experts and country-specific registration practices as part of the general national surveillance purposes. This was done as we anticipated that country-specific definitions were likely better at identifying comorbidity-related risk groups within each country than a common set of code definitions.

**Supplementary Table S3. Specification and emulation of the pragmatic target trial of vaccination with a monovalent XBB.1.5-containing covid-19 mRNA vaccine and risk of severe covid-19 outcomes using nationwide-based registry data across three Nordic countries.**

| Protocol | Target Trial Specification | Target Trial Emulation |
|---|---|---|
| **Eligibility criteria** | • Aged ≥65 years in Denmark, Finland, and Sweden at start of study period (1 October 2023)<br>• Have a known residency within the specific country at start of study period<br>• Have received ≥four covid-19 vaccine doses (of AZD1222 and/or [original/BA4-5/BA-1 bivalent] mRNA vaccines [AZD1222 as part of the primary vaccination course only]) prior to the start of study period<br>• No history of Covid-19 hospitalization prior to the start of study period | Same as for the target trial. |
| **Treatment strategies** | 1) Receive a monovalent XBB.1.5-containing covid-19 vaccine at baseline and do not receive additional covid-19 vaccine doses during follow-up<br>2) Do not receive a monovalent XBB.1.5-containing covid-19 vaccine at baseline and continue being unvaccinated during follow-up | Same as for the target trial.<br>We define the date of vaccination with the XBB.1.5-containing covid-19 vaccine (that is, the index date) according to the registered date of administration. |
| **Treatment assignment** | Individuals are randomly assigned to a strategy at baseline in a 1:1 ratio | Individuals are assigned to the strategy compatible with their treatment received at that time (XBB.1.5-containing vaccine recipient and non-recipient); randomization is assumed conditional on matching (in a 1:1 ratio) on baseline covariates; vaccine non-recipient are assigned the index date of the matched vaccine recipient. |
| **Outcomes** | - Covid-19 hospitalisation: inpatient hospitalisation with a registered covid-19-related diagnosis and a positive PCR test for SARS-CoV-2 (within 14 days before to 2 days after the day of admission)<br>- Covid-19 death: death within 30 days of a positive PCR test for SARS-CoV-2 | Same as for the target trial. |
| **Follow-up** | Follow-up for each individual will start at day 8 from treatment assignment (to ensure full immunisation among XBB.1.5-containing covid-19 vaccine recipients) and end on day of outcome event, weeks 12 has passed, death, emigration, or end of the study period (29 February 2024), whichever occurs first. | Same as for the target trial. |
| **Causal contrast of interest** | Per-protocol | Observational analogue to per-protocol effect. |
| **Statistical analysis** | The Aalen-Johansen estimator will be used to obtain cumulative incidence for each treatment strategy during follow-up (with death as a competing risk). The cumulative incidence across treatment strategies are used to calculate risk ratios (to obtain comparative vaccine effectiveness) and risk differences at the end of week 12. In addition, person-time since baseline will be stratified by consecutive 3-week intervals to estimate changes in comparative vaccine effectiveness per 3 weeks of follow-up. Subgroup analyses by sex (female/male), age (65-74/≥75 years), number of covid-19 vaccine doses received, and seasonal influenza vaccination status. | Same as for the target trial except observational analogues of per-protocol. |

**Supplementary Table S4. Covid-19 hospitalisation and death outcome definitions.**

| **Outcome variable** | **Country** | **Data source and details** |
|---|---|---|
| Covid-19 hospitalisation | Denmark | *The National Patient Register and the Danish Microbiology Database.* Defined as a hospitalization with a PCR positive test for SARS-CoV-2 within 14 days before to 2 days after the admission date, b) inpatient contact or at least 12 hours of contact, and c) a covid-19 relevant diagnosis code (ICD-10: B342, B342A, B948A, B972, B972A, B972B, B972B1, Z038PA1) |
| | Finland | *National Care Register for Health Care and the National Infectious Diseases Register.* Defined as a hospitalization with a PCR positive test for SARS-CoV-2 within 14 days before to 2 days after the admission date, b) inpatient hospital contact, and c) a covid-19 relevant main diagnosis (ICD-10: J00-J22, J46, J80-J84, J851, J86, U071, U072). |
| | Sweden | *The Swedish Patient Register and the Register on surveillance of notifiable communicable diseases (SmiNet).* Defined as a hospitalization with a PCR positive test for SARS-CoV-2 within 14 days before to 2 days after the admission date, b) inpatient contact or at least 12 hours of contact, and c) a covid-19 relevant diagnosis code (ICD-10: U071, U072, U109) |
| Covid-19 death | Denmark | *The Civil Registration System and the Danish Microbiology Database.* Defined as (the date of) death within 30 days after PCR positive test for SARS-CoV-2. |
| | Finland | *The Finnish Population Information System and the National Infectious Diseases Register. D*efined as (the date of) death within 30 days after PCR positive test for SARS-CoV-2. |
| | Sweden | *The Total Population Register, the Cause of Death Register, and the Swedish Patient Register and the Register on surveillance of notifiable communicable diseases (SmiNet).* Defined as (the date of) death within 30 days after PCR positive test for SARS-CoV-2. |

**Supplementary Figure S1. Graphical illustration of the study design.**

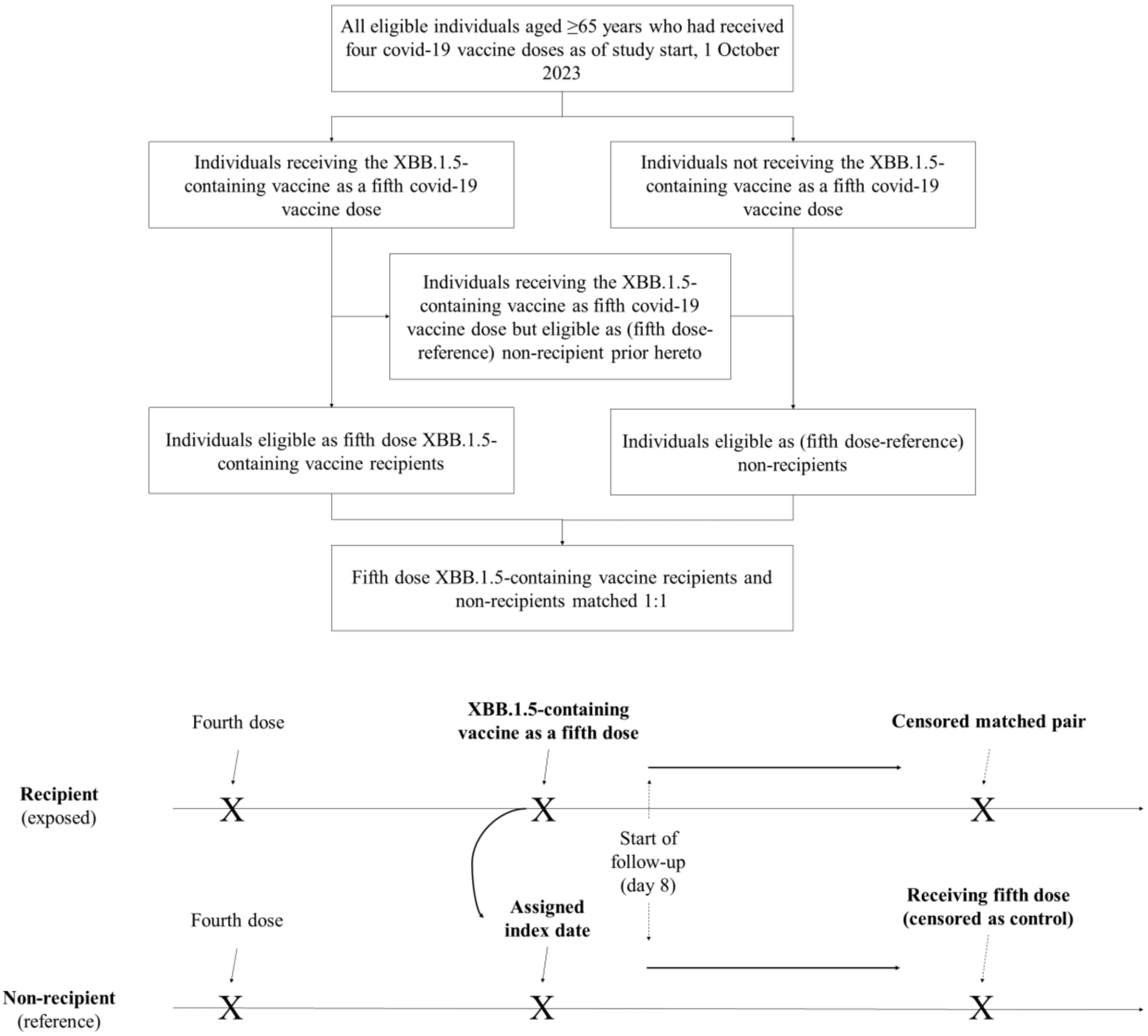

Figure illustrates the study design and uses the example of the XBB.1.5-containing vaccine being received as a fifth covid-19 vaccine dose. By study design, the XBB.1.5-containing vaccine could be received as a ≥fifth covid-19 vaccine dose as we included individuals who had received at least four covid-19 vaccine doses prior to start of the study period. XBB.1.5-containing vaccine recipients were matched 1:1 exact without replacement to (at that calendar date) non-recipients who had received the same number of covid-19 vaccine doses prior to study start on age, calendar time of last prior covid-19 vaccine dose received (in monthly bins; e.g., month of receiving the 4th dose for matched pairs where the XBB.1.5-containing vaccine was administered as a 5th dose), sex, region of residence, vaccination priority groups, and number of selected comorbidities (by 0, 1, 2, or ≥3 of chronic pulmonary disease, cardiovascular conditions, diabetes, autoimmunity-related conditions, cancer, and moderate-to-severe renal disease); see supplementary table S2 for definition details.

**Supplementary Figure S2. Density plots of the distribution of age and index date across countries.**

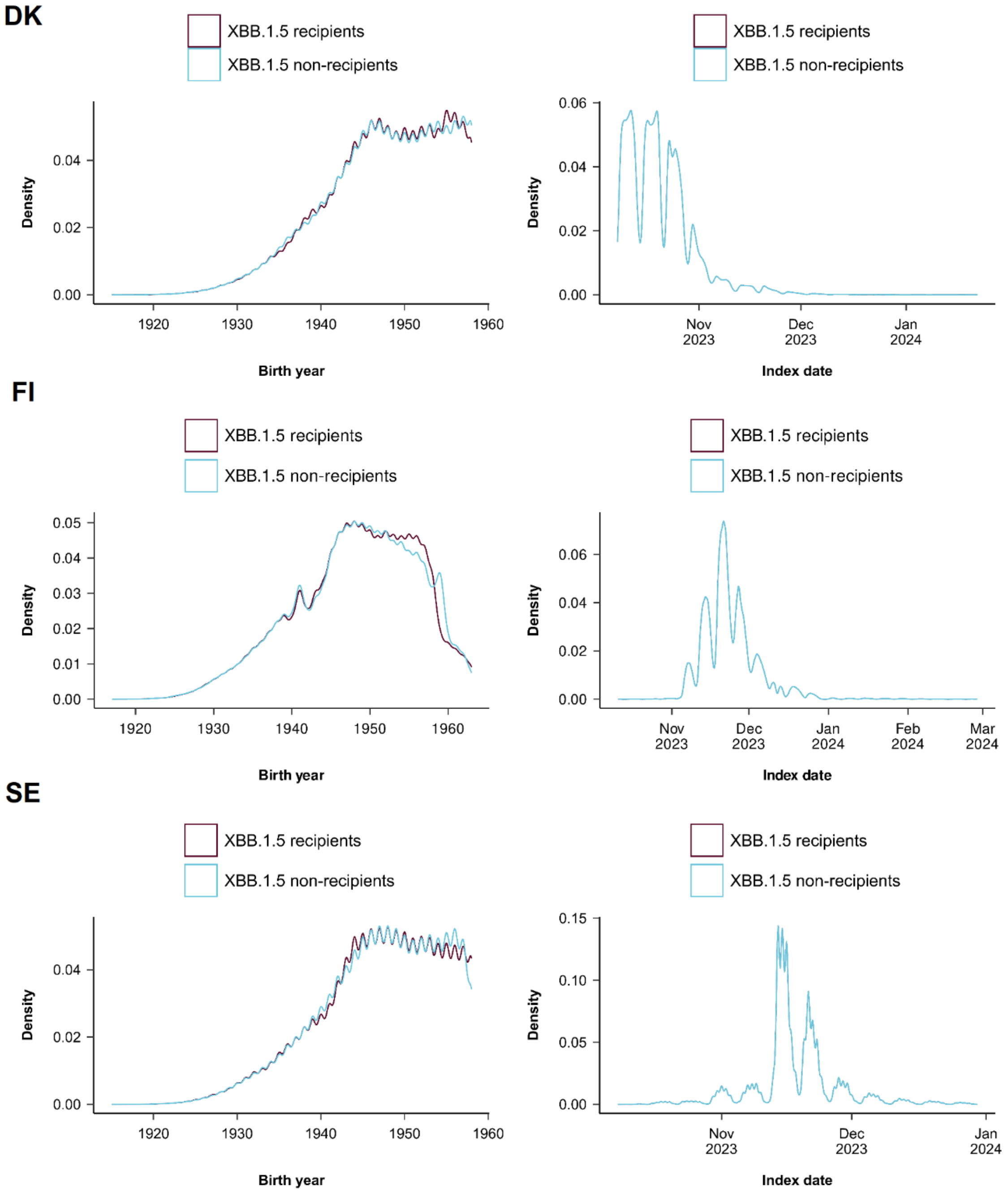

DK denotes Denmark, FI Finland, SE Sweden, and XBB.1.5 XBB.1.5-containing vaccine.

**Supplementary Table S5. Sensitivity analysis of risk of hospital admission and death related to covid-19 comparing XBB.1.5-containing vaccine recipients with non-recipients in Denmark, Finland and Sweden, 1 October 2023 to 29 February 2024, starting follow-up 21 days of the vaccination date.**

| Covid-19 outcomes | Contributing countries | Events / person-years: XBB.1.5-containing vaccine recipients | Events / person-years: XBB.1.5-containing vaccine non-recipients | Risk difference (95% CI) per 100,000 individuals | Comparative vaccine effectiveness (95% CI), % |
|---|---|---|---|---|---|
| Hospital admission | DK, FI | 286/92,640 | 836/91,727 | -137.0 (-385.1 to 111.2) | 57.6 (29.8 to 85.5) |
| Death | DK, FI, SE | 212/149,004 | 879/147,628 | -92.2 (-118.3 to -66.2) | 76.2 (64.2 to 88.2) |

CI denotes confidence interval, DK Denmark, FI Finland, and SE Sweden.

**Supplementary Table S6. Risk of hospital admission and death related to covid-19 at 6 weeks of follow-up comparing XBB.1.5-containing vaccine recipients with non-recipients in Denmark, Finland and Sweden, 1 October 2023 to 29 February 2024.[a]**

| | | Events / person-years | | | |
|---|---|---|---|---|---|
| **Covid-19 outcomes** | **Contributing countries** | **XBB.1.5-containing vaccine recipients** | **XBB.1.5-containing vaccine non-recipients** | **Risk difference (95% CI) per 100,000 individuals** | **Comparative vaccine effectiveness (95% CI), %** |
| Hospital admission | DK, FI, SE | 787/120,264 | 2207/119,748 | -127.2 (-204.4 to -50.0) | 64.1 (49.1 to 79.1) |
| Death | DK, FI, SE | 202/125,511 | 1022/125,065 | -73.4 (-103.0 to -43.8) | 83.0 (74.1 to 91.8) |

CI denotes confidence interval, DK Denmark, FI Finland, and SE Sweden. [a]Individuals were followed for 6 weeks (from 1 week after the vaccination date).

### Supplementary References.

1. Schmidt M, Pedersen L, Sørensen HT. The Danish Civil Registration System as a tool in epidemiology. Eur J Epidemiol 2014;29(8):541–9.

2. Krause TG, Jakobsen S, Haarh M, Mølbak K. The Danish vaccination register. Euro Surveill 2012;17(17):20155.

3. Voldstedlund M, Haarh M, Mølbak K, MiBa Board of Representatives. The Danish Microbiology Database (MiBa) 2010 to 2013. Euro Surveill 2014;19(1):20667.

4. Schmidt M, Schmidt SAJ, Sandegaard JL, Ehrenstein V, Pedersen L, Sørensen HT. The Danish National Patient Registry: a review of content, data quality, and research potential. Clinical Epidemiology 2015;449.
